# Multi-Modal Sleep Measurement and Alignment Analysis in Outpatients with Major Depressive Episode

**DOI:** 10.1101/2025.04.29.25326308

**Authors:** Afrooz Mahir, Nguyen Luong, Ilya Baryshnikov, Annasofia Martikkala, Erkki Isometsä, Talayeh Aledavood

## Abstract

**Study Objectives:** Sleep plays a crucial role for mental health. This study examines sleep tracking in naturalistic settings for patients with major depressive episodes (MDE) using actigraphy, smartphone data, bed sensors, and the ecological momentary assessment (EMA) and assesses discrepancies between these modalities.

**Methods:** We measured sleep onset, offset, and total sleep time (TST) over two weeks for 172 participants, including healthy controls and three MDE subgroups (borderline personality disorder, major depressive disorder, and bipolar disorder). Agreement between measurement modalities was assessed using Bland-Altman plots and Pearson correlation. Predictors of sleep alignment were analyzed using mixed-effects models, accounting for demographics, daylight length, and participant subgroup.

**Results:** Patients showed greater sleep variability than healthy controls. Actigraphy overestimated TST compared to bed sensors (0.48 min) and smartphones (0.99 min), while the smartphone underestimated TST compared to other modalities. Older age improved alignment between actigraphy and bed sensors, as well as smartphone and bed sensor sleep offset. TST alignment (smartphone vs. bed sensor) was worse in females and bipolar/borderline patients. Longer daylight duration improved TST and sleep offset alignment across modalities.

**Conclusions:** Our study highlights measurement biases, seasonal effects, and demographic factors associated with discrepancies in objective sleep measures. While these modalities show potential and offer several advantages in assessing sleep over longer periods, the discrepancies and factors associated with misalignment should be considered in future studies or clinical settings.

**Statement of Significance:** Tracking sleep in psychiatric patients is challenging due to frequent sleep disturbances, making accurate assessment crucial for diagnosis and care. Traditional methods are limited to lab settings, restricting long-term monitoring. This study evaluates the alignment of naturalistic sleep tracking using actigraphy, bed sensors, smartphone data, and self-reports in both healthy individuals and patients with depressive disorders. Our findings demonstrate the feasibility of using these non-invasive methods to monitor sleep for patients with major depressive episodes. We uncover systematic biases in sleep estimates across modalities, reveal demographic and environmental factors that influence measurement agreement, and show that psychiatric populations exhibit more variability in sleep patterns. This work addresses a critical gap in validating consumer-grade sleep tracking technologies for psychiatric populations in naturalistic contexts.

## 1 Introduction

Sleep is a fundamental aspect of mental and physical well-being, playing a crucial role in emotional regulation, cognitive function, and overall health,^1,2.3^ Insufficient and excessive sleep are both linked to cognitive impairment, heightened stress, and increased risk of chronic diseases.^4–6^ In psychiatric populations, sleep abnormalities are especially pronounced, with insomnia and hypersomnia being core symptoms of major depressive disorder (MDD),^7^ and hypersomnia is frequently observed in bipolar disorder (BD).^8^ Moreover, deviations from typical sleep are associated with elevated suicide risk, mood instability, and early relapse indicators in many psychiatric disorders, including mood disorders.^9–11^ Given these profound implications, accurately capturing sleep patterns is essential for psychiatric assessments. However, sleep is typically measured in laboratory settings, which limits its applicability for real-world monitoring. This limitation makes obtaining reliable long-term measurements challenging.

Polysomnography (PSG) is the gold standard for sleep assessment but is typically limited to controlled environments.^12–14^ Alternative methods, such as sleep diaries and questionnaires, offer greater flexibility but are prone to recall bias,^15^ often leading to discrepancies between subjective reports and objective measurements.^16, 17^

Wearable technology, such as actigraphy, provides a more practical solution with reasonable accuracy in tracking some sleep parameters, such as onset and offset.^18^ However, actigraphy has limitations, including potential discomfort and the inability to measure physiological markers like heart rate and respiration.^19^ More recent advancements in sleep monitoring have introduced contactless methods, such as ballistocardiography (BCG)-based bed sensors and smartphone-based sleep tracking.^20^ While these newer modalities show promise, their validity and comparability, especially in assessing patients with depressive disorders in real-world settings, remain underexplored. To address this gap, we employ a multi-modal approach, combining actigraphy, bed sensors, smartphone-based tracking, and the ecological momentary assessment (EMA)^21^ to assess sleep.

A crucial issue in sleep research is the discrepancy between different measurement modalities. Studies comparing PSG with actigraphy and self-reports have consistently shown variation in onset and offset measurements.^13, 14^ The divergence is particularly pronounced in psychiatric populations, where sleep misperception, the tendency to perceive one’s sleep inaccurately, is common.^15, 22^ While actigraphy has been validated against PSG, the role of newer modalities like bed sensors and smartphone data requires further investigation.^23^

Beyond measurement discrepancies, demographic, psychological, and environmental factors introduce additional variability. For example, chronotype, gender, and age, as well as external variables such as seasons, are associated with sleep patterns,^24, 25^ with evening chronotypes displaying a higher risk of depressive symptoms^26^ and women generally reporting longer sleep.^27^ Psychological conditions such as depression further complicate measurement accuracy due to circadian rhythm disruptions.^28^ Moreover, environmental influences, including artificial light exposure and daylight saving time transitions, can also distort sleep assessment.^29–31^

Given these complexities, this study investigates how sleep onset, offset, and total sleep time (TST) measurements compare across actigraphy, smartphone data, bed sensors, and EMA. Additionally, it examines whether these measurements differ between healthy controls and patients with depression, providing insights into the extent of alignment in sleep assessments. Finally, this research explores the demographic, psychological, and environmental factors contributing to discrepancies between measurement modalities, highlighting the sources of error and bias. By addressing these questions, this study contributes to the area of naturalistic sleep research, offering a deeper understanding of sleep monitoring technologies and their implications for psychiatric evaluation, monitoring, and intervention.

## 2 Method

### 2.1 Data collection

The MoMo-Mood study^32^ explored the effectiveness of wearable technology in monitoring sleep and mood in individuals with psychiatric disorders as well as finding behavioural markers of depression from passively collected data.^32, 33^ The study included 164 participants, of which 133 were diagnosed with a current Major Depressive Episode (MDE), including 85 patients with MDD, 27 with Borderline Personality Disorder (BPD), and 21 with BD, alongside 31 healthy controls. Additionally, a pilot study was conducted with 37 participants, including 14 MDD patients and 23 healthy controls. The main and the pilot studies had a similar design and data from both studies were included in the analysis. The study period was divided into two phases: an initial two-week active phase and a subsequent passive phase lasting up to one year. In this work, we focused exclusively on the active phase of data collection. During this phase, participants wore wrist-worn actigraphy devices and used bed sensors to monitor sleep patterns. Concurrently, smartphone data was gathered through the AWARE application,^34^ which tracked various behavioural metrics.^32^ Participants were also prompted five times daily to report on their mood, energy levels, and other psychological states using a 7-point Likert scale. This analysis did not include the passive phase, which involved passive smartphone monitoring and bi-weekly assessments of the Patient Health Questionnaire (PHQ-9). A more detailed description of the questions can be found elsewhere.^35^

### 2.2 Sleep and activity monitoring protocol

Participants used wearable devices, specifically Philips Actiwatch 2 actigraphs, to measure activity levels and sleep for two weeks without recharging. This ensured that participants did not have to remove the device during the monitoring period. They wore the actigraph on their wrist throughout the active phase, ensuring continuous data collection. They were instructed to remove the device only during sauna use, which is a common activity in Finland.

In addition to the actigraphs, nearable devices, including bed sensors and smartphones, were also used for sleep monitoring. The Murata SCA11H nodes, based on ballistocardiography technology, were used for bed sensing. Each participant received a pre-configured WiFi router to enable automatic data transfer to the study server. The participants were asked to place the Murata SCA11H node under their mattress, positioned close to them but not directly beneath them. Alternatively, they could be attached to the bed frame near the participant. The sensors processed information on-site and produced metrics such as pulse rate, heart rhythm variability, breathing rate, cardiac output, and signal intensity at a 1 Hz rate.^36^

In this study, smartphones were also used for sleep tracking due to their non-invasive nature and ability to monitor behaviours.^20^ We collected communication timestamps (calls and texts), anonymous contact identifiers, smartphone screen activity events (screen on, off, lock, unlock), location data, app usage patterns, and battery state. In this study, we only analysed screen activity events to identify sleep periods.

Furthermore, smartphones facilitated the use of EMA, which was employed to gather real-time data on sleep and participants’ emotional states through-out the study.^32^ During the active phase, participants received multiple EMA prompts daily on their smartphones. These included a morning questionnaire to assess sleep from the previous night and an evening questionnaire to track the activities of the day. Additionally, three randomly timed prompts were sent throughout the afternoon within specific time ranges. This schedule allowed for collecting subjective data on sleep, mood, energy levels, physical activity, and other psychological factors.^35^ Data from all modalities were gathered through the Niima data collection platform^37^ and was pre-processed with the Niimpy behavior analysis toolbox.^38^

### 2.3 Device data export procedures and analysis

Sleep data was derived using proprietary algorithms from both the actigraph and bed sensor, which were then transformed into sleep parameters for analysis. For the actigraph, raw data was aggregated into 30-second intervals, with each interval assigned one of three possible labels based on activity levels. The ‘ACTIVE’ label indicated high activity, indicating the user was awake and moving. The ‘REST’ label represented low activity, suggesting the user was resting but not necessarily asleep. The ‘REST-S’ label denoted sustained low activity, indicating that the user was likely asleep. To identify sleep episodes, the data was sorted chronologically, and continuous REST-S intervals were grouped together and treated as a single sleep episode.

The bed sensor collected raw data on heart rate and respiratory rate at 1-second intervals. This data was processed using the manufacturer’s algorithm to generate labels indicating bed occupancy and physiological states.^32^ Sleep periods were identified based on the status variable, where a status of ‘1’ indicated sleep, and ‘0’, ‘2’, and ‘3’ represented non-sleep states (e.g., not in bed, awake in bed, or signal overload). Continuous status ‘1’ intervals were treated as sleep episodes, grouped together as part of the same sleep period.

For the smartphone data, sleep was inferred based on the lock and unlock status as indicators of inactivity, following the procedure described in^32^ and used in previous studies.^39, 40^ Periods of inactivity, identified when the device was locked, were assessed to determine sleep periods. The longest stretch of inactivity was classified as nocturnal sleep. The lock/unlock status was converted into a binary format, and the longest inactivity periods were tracked to calculate sleep onset, sleep offset, and TST.

In addition, the EMA questions and responses, initially in Finnish, were translated into English. Data was selected based on responses to the question, “How many hours did you sleep last night?” to examine the relationship between subjective and sensor-based sleep data. Categorical TST ranges were transformed into numeric values (e.g., “6-7 hours” was converted to 6.5 hours). Data preprocessing was performed on the smartphone, actigraph, and bed sensor data. First, missing data was removed to ensure completeness. Second, timestamps were standardized to the Europe/Helsinki time zone to maintain consistency across all data sources. Third, sleep data was aligned using a 15:00–15:00 time frame, meaning each 24-hour period started and ended at 15:00 each day. This approach was chosen to account for variations in sleep schedules and to standardize sleep cycle measurements across participants.

A five-minute threshold was applied to identify sleep periods, allowing gaps of up to five minutes between consecutive sleep intervals to be treated as part of the same sleep episode. TST outside the 3–13 hour range was excluded, following established guidelines.^41^ This threshold was set to exclude implausible TST values, as unusually short or long sleep episodes could indicate data errors or atypical patterns.

Finally, sleep onset and offset were determined by identifying the start and end times of the longest detected sleep episode within each 24-hour period. Participants with missing values for these parameters were excluded from the analysis. Figure 1 illustrates how sleep periods were identified using the sleep labels provided by the manufacturer for both the actigraph and bed sensor.

**Figure 1.**
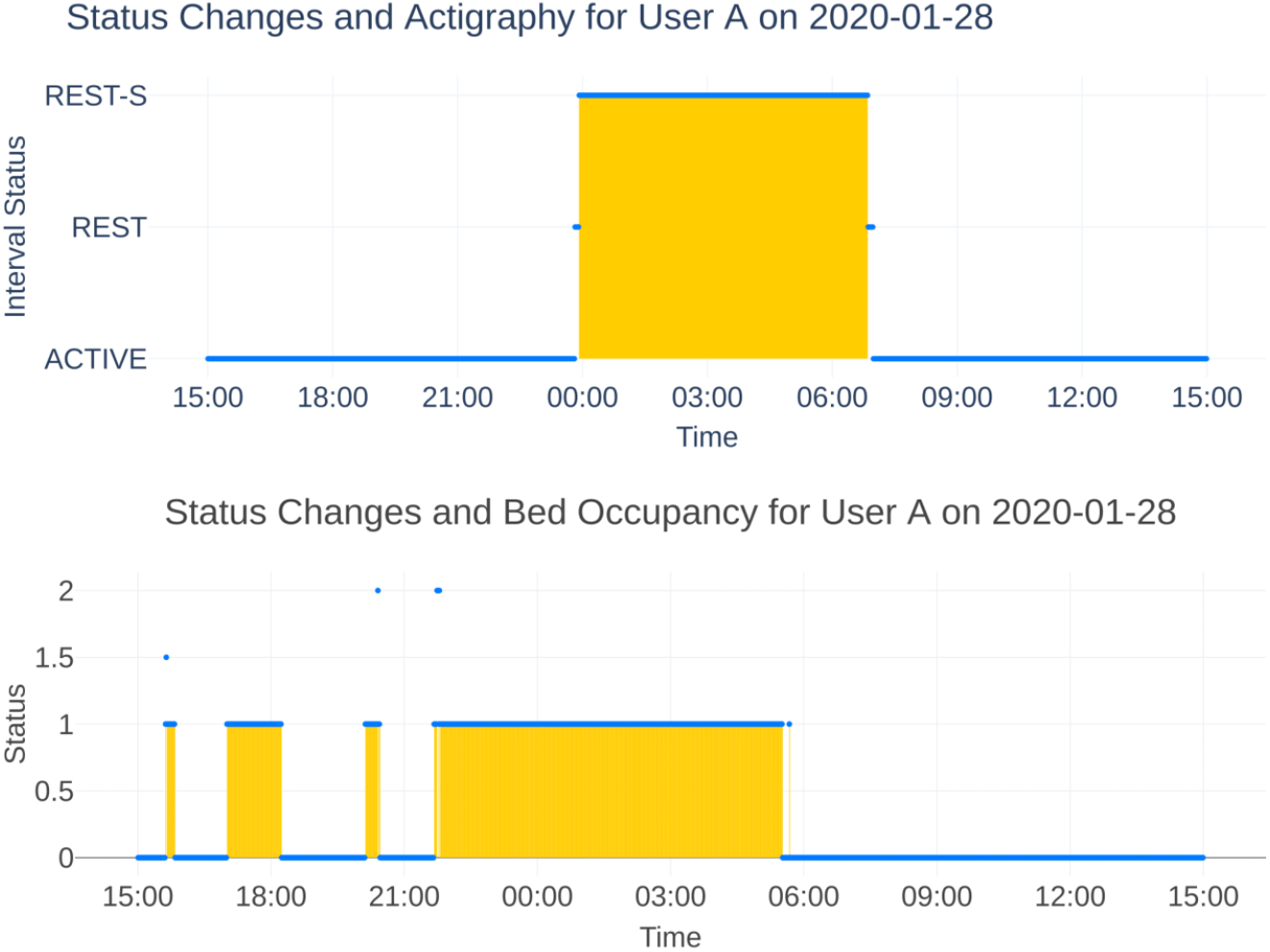
Status changes identified by proprietary algorithms of the actigraph and bed sensor. For the actigraph, the sleep period began at 23:55 and ended at 06:51, marked by “Rest-S,” while the bed sensor indicated “Status 1,” showing the bed was occupied and the user was asleep from 21:40 to 05:30. This displays differences in sleep period detection and wakefulness between the two modalities.

### 2.4 Missing data and outlier handling procedures

Although 164 participants were initially recruited for the main study, 13 participants (7.9%) did not submit passive data and were removed from all evaluations. Additionally, missing data resulted from various factors, including device non-compliance, such as failing to wear or charge the device, user dropout, inconsistent usage, and data cleaning procedures. Technical issues, such as battery depletion, sensor malfunctions, or connectivity failures, may have also contributed to data loss. After filtering out missing data, the sample included 173 unique users from both the main and pilot studies. The datasets included 102 actigraph users, 108 bed sensor users, and 148 smartphone users.

Additionally, the two largest outliers from both the onset and offset columns were identified and removed. Due to the overlap between the outliers in both columns, three outliers were removed from the bed sensor and actigraph datasets and two from the smartphone dataset, resulting in the exclusion of eight users in total. Manual checks revealed that these outliers were due to device-related issues, including improper device wear for the actigraph, placement or configuration errors for the bed sensor, and technical problems with the smartphone data. Following data cleaning, the final sample size reduced to 172 users, and were as follows: actigraph (29 healthy controls, 70 patients; total N = 99), bed sensor (34 healthy controls, 71 patients; total N = 105), smartphone (46 healthy controls, 100 patients; total N = 146), and EMA (48 healthy controls, 109 patients; total N = 157)

### 2.5 Statistical analysis

To assess the agreement between different sleep measurement modalities, Bland-Altman plots^42^ were created by plotting the differences between paired measurements against their mean. These plots help visualize the differences between two modalities, revealing any consistent bias and the range where the measurements align. Sleep onset, offset, and TST comparisons were made across modalities using this approach, with 95% limits of agreement (LoA) calculated. To account for day-to-day variability, data were aggregated by averaging user data across multiple days. Pearson’s correlation analysis was conducted to explore the relationships between sleep parameters from different modalities. Normality was first assessed using the Shapiro-Wilk test.^43^ If slight deviations from normality were observed, QQ plots were used for cross-checking,^44^ which ultimately confirmed normality. The size of the correlation coefficients was interpreted based on the thresholds from.^45^ A high correlation ranged from 0.70 to 1.00, a moderate correlation ranged from 0.50 to 0.70, a low correlation was between 0.30 to 0.50, and a negligible correlation was considered when ranged from 0.00 to 0.30. Finally, 95% confidence intervals were calculated for the regression slopes. Finally, linear mixed models^46, 47^ were employed to investigate factors associated with the alignment of sleep parameters between modalities. The alignment of sleep parameters between measurement modality pairs was used as the dependent variables, represented as the absolute difference between the two modalities. The models adjusted for demographic factors, including age and gender. Chronotype was included as a covariate, measured by the Morning-Eveningness Questionnaire (MEQ) and categorized on a five-point scale: definitely morning type (70–86), moderately morning type (59–69), neither type (42–58), moderately evening type (31–41), and definitely evening type (16–30). Given the unique geographical position of Finland, the seasonal factor was controlled for using day length duration. Between-group differences were accounted for by comparing three patient groups against using the healthy control group as a reference. The model was performed in R Statistical Software^48^ version 4.3.1 using the ‘lme4’ package.^49^ All other analyses were performed in Python 3.12.^50^

## 3 Results

### 3.1 Descriptive Statistics

The demographic characteristics of the participants included in the study (N = 169) are outlined in Table 1. Three users from the pilot study did not provide information on age, gender, or neither; thus, their data was excluded from further analyses. The participants had a mean age of 35.07 years (SD = 12.81), with 73.96% identifying as female. The mean Morningness-Eveningness Questionnaire (MEQ) score was 39.15 (SD = 5.15), reflecting individual chronotype tendencies. Regarding group distribution, 46.15% of the participants were diagnosed with MDD, 29.59% were in the control group, 12.43% with BD, and 11.83% with BPD.

**Table 1:**
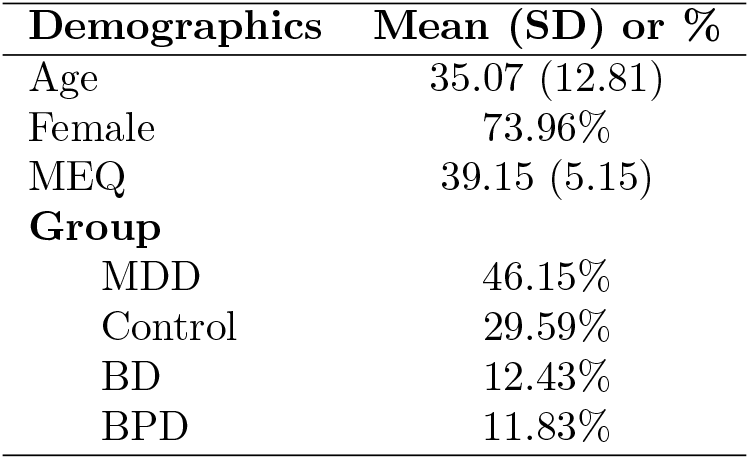
Demographic characteristics of participants (N=169).

| **Demographics** | **Mean (SD) or %** |
| --- | --- |
| Age | 35.07 (12.81) |
| Female | 73.96% |
| MEQ | 39.15 (5.15) |
| **Group** |  |
| MDD | 46.15% |
| Control | 29.59% |
| BD | 12.43% |
| BPD | 11.83% |

Descriptive statistics for sleep onset, offset, and TST across different sleep assessment modalities are presented in Table 2. In the control group, sleep onset times differ. The actigraph recorded the earliest mean onset at 23:48 (SD = 70 min), while the smartphone reported the latest mean onset at 00:01 (SD = 106 min). The bed sensor recorded a mean onset time of 23:58 (SD = 67 min), falling between the actigraph and the smartphone. In the patient group, mean onset times were generally later, with the bed sensor recording the earliest mean onset at 00:19 (SD = 100 min) and the actigraph detecting the latest at 00:35 (SD = 92 min). The smartphone recorded a mean onset time of 00:30 (SD = 101 min), with slight variations across groups.

**Table 2:** Mean sleep parameters (onset, offset, and total sleep time) and SD across different assessment methods for control and patient groups.

|  |  | **Bed Sensor** | **Actigraph** | **Phone** | **ESM** |
| --- | --- | --- | --- | --- | --- |
| **Onset** | Control | 23:58 ± 67 m | 23:48 ± 70 m | 00:01 ± 106 m | - |
| | Patients | 00:19 ± 100 m | 00:35 ± 92 m | 00:30 ± 101 m | - |
| **Offset** | Control | 07:33 ± 87 m | 07:44 ± 74 m | 07:06 ± 72 m | - |
| | Patients | 08:24 ± 119 m | 08:58 ± 91 m | 07:32 ± 92 m | - |
| **TST** | Control | 7h 35m ± 78m | 7h 55m ± 62m | 7h 10m ± 87m | 7h 14m ± 35m |
| | Patients | 8h 5m ± 107m | 8h 22m ± 60m | 7h 7m ± 78m | 7h 26m ± 68m |

Patients show later sleep onset times and later sleep offset times compared to controls, with TST generally higher for patients across all measurement methods. Notably, TST recorded by actigraphy was higher than other devices, while the phone-based method recorded the shortest TST.

Sleep offset showed a similar pattern, with differences between groups and measurement modalities. In the control group, the smartphone reported the earliest mean offset at 07:06 (SD = 72 min), while the actigraph recorded the latest mean offset at 07:44 (SD = 74 min). The bed sensor estimated a mean offset of 07:33 (SD = 87 min). In the patient group, offsets occur later, with the smartphone recording the earliest mean offset time at 07:32 (SD = 92 min) and the actigraph identifying the latest at 08:58 (SD = 91 min). The bed sensor reported a mean offset of 08:24 (SD = 119 min), indicating a trend of delayed offset times among patients compared to controls. Furthermore, patients show greater variability in sleep offset times, suggesting more inconsistency in their sleep patterns.

TST is evaluated using both objective measurements and subjective reports from the EMA questionnaires. In the control group, the smartphone recorded the shortest mean TST at 7 hours 10 minutes (SD = 87 min), while the actigraph recorded the longest at 7 hours 55 minutes (SD = 62 min). The bed sensor estimated a mean TST of 7 hours 35 minutes (SD = 78 min), and subjective reports from the EMA questionnaires indicated a mean of 7 hours 14 minutes (SD = 35 min). For the patient group, the mean TST is generally longer, with the smartphone recording the shortest mean TST at 7 hours 7 minutes (SD = 78 min) and the actigraph recording the longest at 8 hours 22 minutes (SD = 60 min). The bed sensor estimated a mean TST of 8 hours 5 minutes (SD = 107 min), and EMA responses showed a mean of 7 hours 26 minutes (SD = 68 min). Overall, these results show relatively consistent trends across the sleep modalities.

### 3.2 Alignment of Sleep Parameters Across Assessment Modalities Sleep Onset

Bland-Altman plots comparing sleep onset times across modalities are presented in the figure 2. The mean difference between sleep onset was not statistically significant (Table 3). However, sleep onset showed significant positive correlations across modalities, as displayed in Figure 3. Actigraph and bed sensor onset times had the strongest correlation (r = 0.70, *p <* 0.001, 95% CI [0.58, 0.96]), indicating a high level of agreement between these modalities. The correlation between actigraph and smartphone onset times was moderate (r = 0.69, *p <* 0.001, 95% CI [0.60, 0.95]), indicating that these two modalities capture similar sleep onset patterns. The correlation between smartphone and bed sensor onset times was also moderate (r = 0.50, *p <* 0.001, 95% CI [0.32, 0.69]), suggesting greater variability in smartphone-based measurements.

**Table 3:** Mean bias and statistical comparison of sleep metrics across assessment methods.

| | Comparison | Mean bias (min) ± SD (min) | t (p) |
| --- | --- | --- | --- |
| **Onset** | Actigraph vs Bed Sensor | 1.80 ± 61.79 | 0.24 (0.81) |
| | Phone vs Bed Sensor | 6.08 ± 85.93 | -3.35 (0.51) |
| | Actigraph vs Phone | -4.10 ± 74.34 | 6.85 (0.62) |
| **Offset** | Actigraph vs Bed Sensor | 30.39 ± 84.65 | 3.00 (p<0.001) |
| | Phone vs Bed Sensor | -37.86 ± 106.60 | -3.35 (p<0.001) |
| | Actigraph vs Phone | 64.85 ± 86.26 | 6.85 (p<0.001) |
| **TST** | Actigraph vs Bed Sensor | 0.48 ± 1.72 | 1.51 (0.13) |
| | Actigraph vs Phone | 0.99 ± 1.46 | 6.29 (p<0.001) |
| | Actigraph vs ESM | 0.98 ± 1.11 | 5.43 (p<0.001) |
| | ESM vs Bed Sensor | -0.42 ± 1.55 | -2.49 (0.01) |
| | ESM vs Phone | 0.38 ± 1.33 | 1.81 (0.07) |
| | Phone vs Bed Sensor | -0.58 ± 1.86 | -3.57 (p<0.001) |

Mean biases in onset time are not statistically significant, whereas offset biases are. TST differences are mostly significant, except for actigraphy vs. bed sensor, ESM vs. bed sensor, and phone vs. bed sensor comparisons.

**Figure 2.**
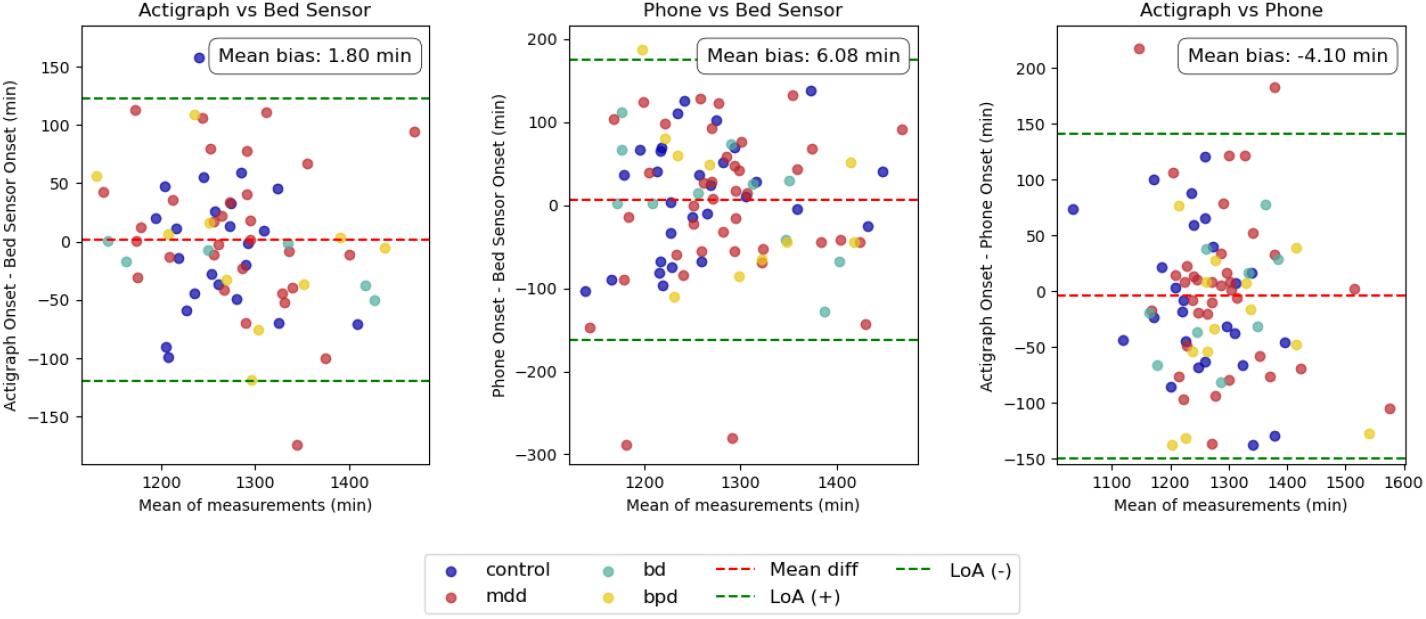
Bland-Altman plot comparing sleep onset times across modalities, with data points colour-coded by group. The red dashed line indicates the mean bias, while the green dashed lines represent the 95% LoA. The smallest mean bias is observed between the actigraph and bed sensor.

**Figure 3.**
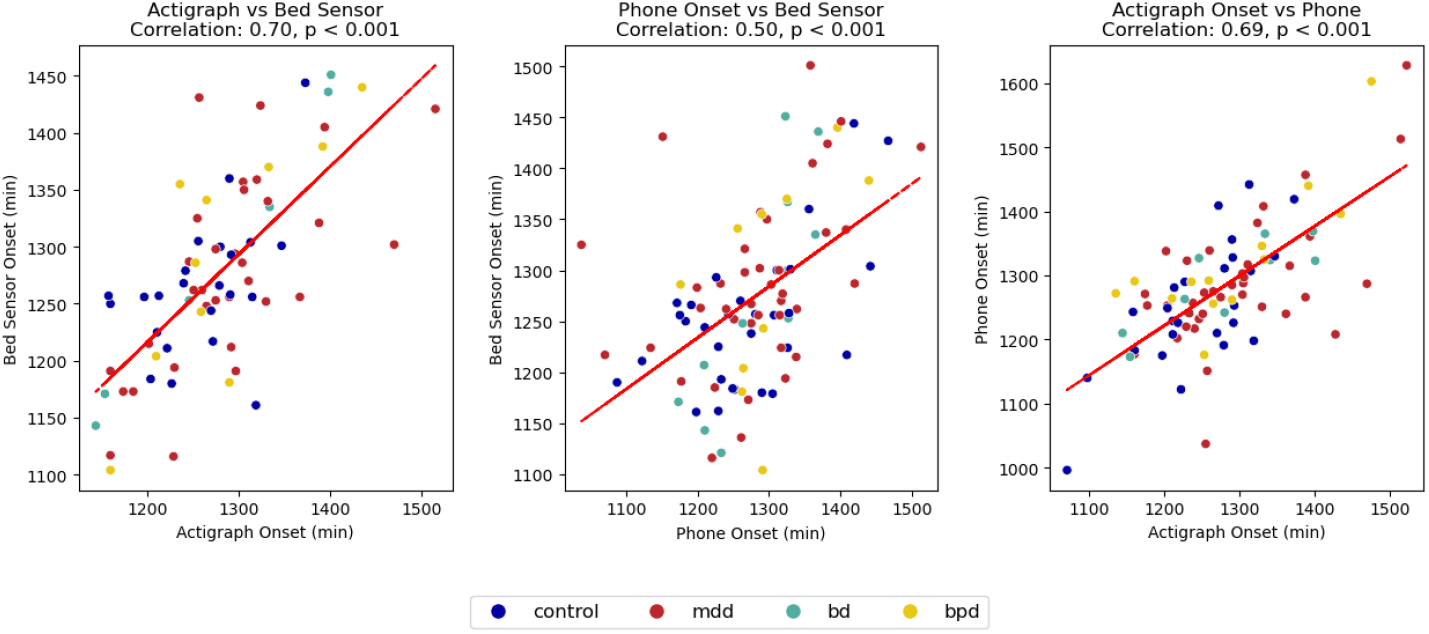
Scatter plots comparing sleep onset times across different modalities, with each point representing a paired data set. Data points are colour-coded to distinguish control and patient groups. All comparisons show positive correlations, with the strongest correlation observed between the actigraph and bed sensor.

### Sleep Offset

Similarly, the comparisons between the offsets recorded by the different modalities are presented in Figure 4. The mean bias for sleep offset across all modalities was statistically significant (*p <* 0.001). In particular, the mean bias between the actigraph and bed sensor offsets was 30.39 minutes (SD = 84.66, 95% CI [10.36, 50.43]), indicating a positive bias, where the actigraph tends to report later onset times compared to the bed sensor. The smartphone offset differed from the bed sensor by a mean of −37.86 min (SD = 106.60, 95% CI [−60.18, −15.53]), showing a negative bias, with the smartphone tending to report earlier onset times compared to the bed sensor. Lastly, the mean difference between the actigraph and smartphone offsets was 64.85 min (SD = 86.26, 95% CI [46.13, 83.57]), reflecting a larger positive bias, with the actigraph reporting significantly later onset times compared to the smartphone.

**Figure 4.**
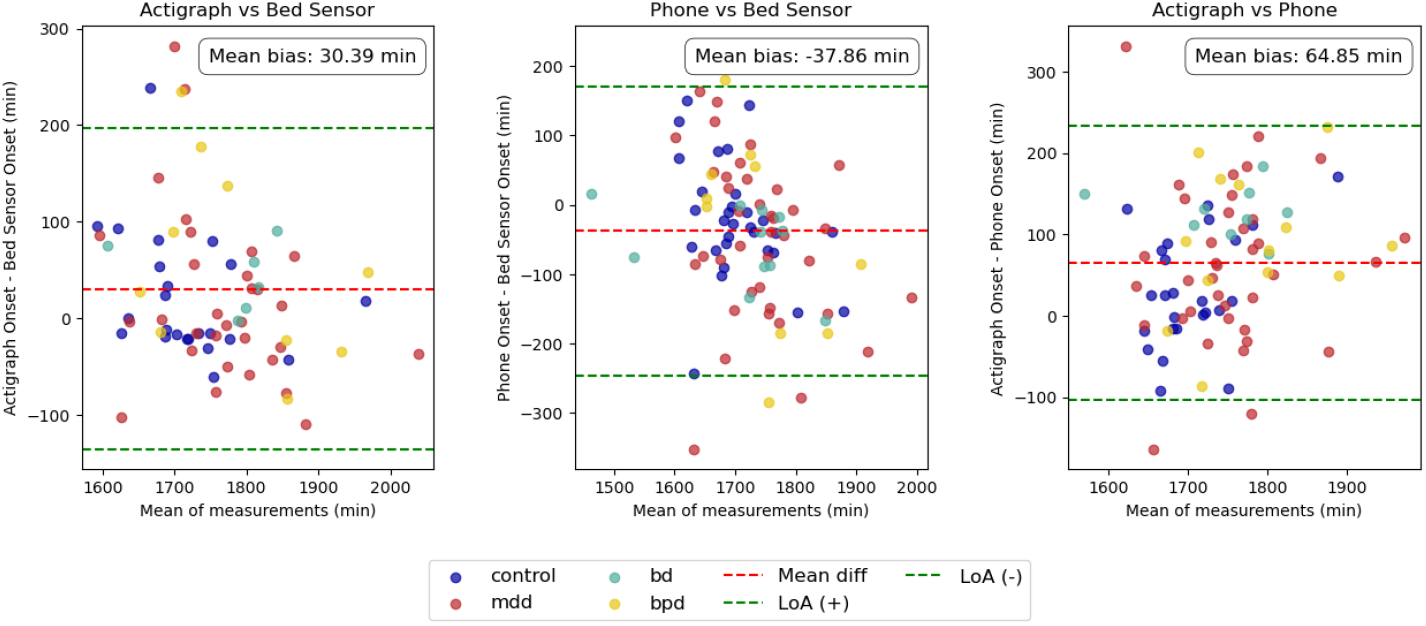
Bland-Altman plot comparing sleep offset times across modalities. The actigraph records later offsets, while the smartphone records earlier offsets relative to other modalities. The largest mean bias in sleep offset is observed between the actigraph and smartphone.

Moreover, all modalities showed moderate to low correlation in measuring sleep offset (Figure 5). The highest correlation was observed between the actigraph and bed sensor (r = 0.65, *p <* 0.001, 95% CI [0.57, 1.01]), followed by the correlation between the actigraph and smartphone (r = 0.51, *p <* 0.001, 95% CI for intercept [0.27, 0.59]). The smartphone and bed sensor showed the lowest correlation (r = 0.43, *p <* 0.001, 95% CI [0.34, 0.88]). Results indicated moderate to low alignment in sleep offset measurements across modalities, with greater variability observed in smartphone-based measurements.

**Figure 5.**
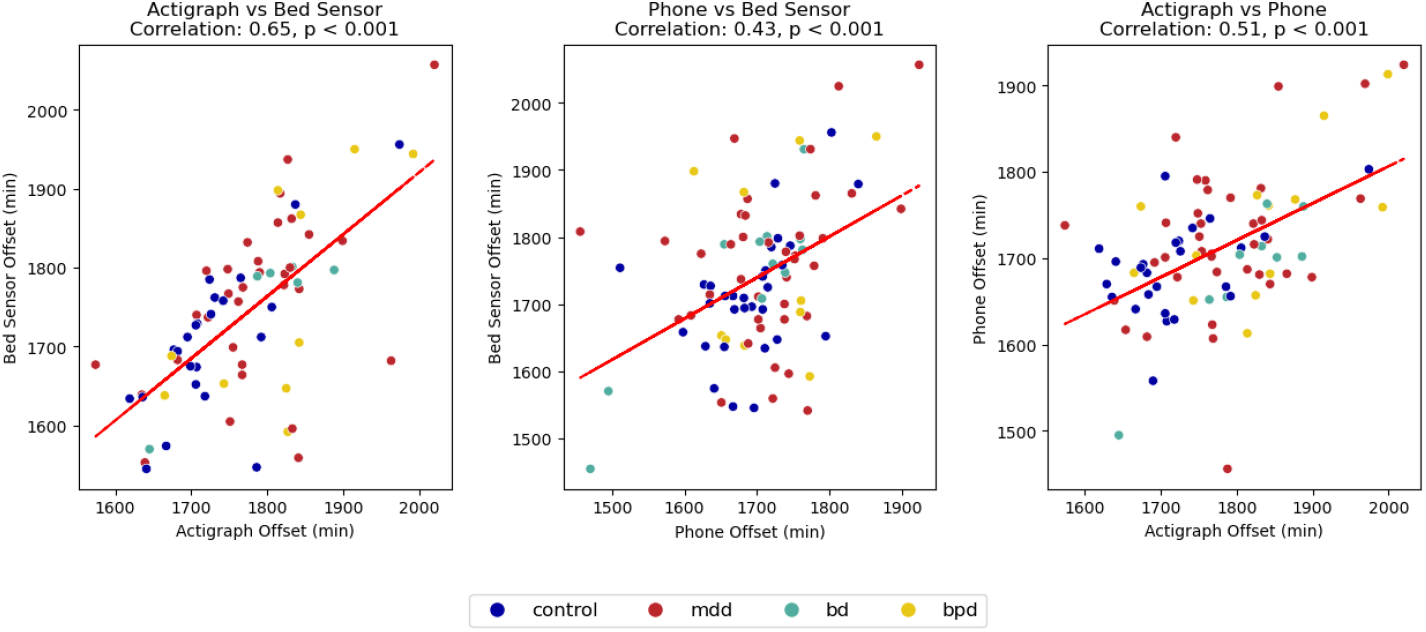
Scatter plots comparing sleep offset times across different modalities. All offsets show a mild positive correlation, with the strongest correlation observed between the actigraph and bed sensor.

### TST

The actigraph consistently reported higher TST compared to the other modalities, as shown in Figure 6. The mean difference between actigraph and bed sensor TST was 0.48 minutes (SD = 1.72, 95% CI [0.18, 0.77]), reflecting a small positive bias, which was not statistically significant (p = 0.13). The actigraph also overestimated TST compared to the smartphone by 0.99 minutes (SD = 1.46, 95% CI [0.74, 1.23]) and by 0.98 minutes (SD = 1.11, 95% CI [0.79, 1.16]) when compared to the EMA data, both of which were statistically significant (*p <* 0.001).

**Figure 6.**
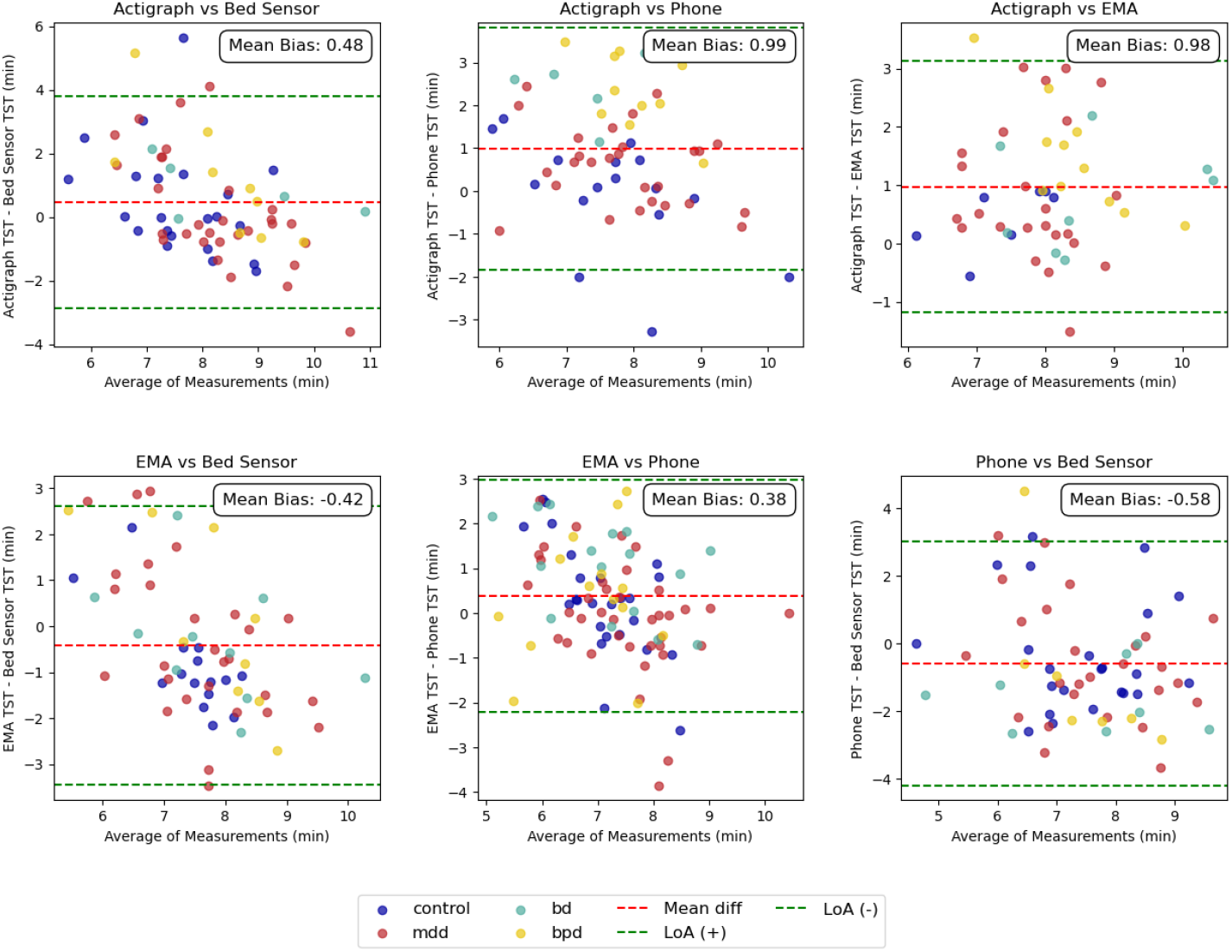
Bland-Altman plot comparing TST across modalities, with data points colour-coded by group. The actigraph reported higher TST compared to other modalities, while the smartphone reported lower values.

EMA slightly underestimated TST relative to the bed sensor and actigraph, but overestimated it compared to the smartphone. The mean difference between EMA and the bed sensor was −0.42 minutes (SD = 1.55 95% CI [−0.67 −0.16]), reflecting a small negative bias, which was statistically significant (p = 0.01). In comparison to the smartphone, EMA had a mean difference of 0.38 minutes (SD = 1.33 95% CI [0.16 0.59]), indicating a very small positive bias, though this difference was not statistically significant (p = 0.07). Lastly, the smartphone underestimated TST compared to the bed sensor, with a mean difference of −0.58 minutes (SD = 1.86, 95% CI [−0.88 −0.27]), which was statistically significant *p <* 0.001. These results suggest that while there are minor differences, the modalities generally agree on TST.

Pearson correlations revealed varying levels of agreement between modalities for TST (Figure 7). The strongest low correlation was observed between actigraph and EMA (r = 0.45, *p <* 0.001, 95% CI [0.20, 0.63]), indicating a stronger alignment between objective and subjective reports. Actigraph and bed sensor sleep measurements demonstrated a low positive correlation (r = 0.30, *p* = 0.011, 95% CI [0.12, 0.91]), while actigraph and smartphone measurements showed a similar low correlation (r = 0.30, *p* = 0.006, 95% CI [0.10, 0.60]). The correlation between EMA and bed sensor data was less pronounced and not statistically significant (r = 0.21, *p* = 0.080, 95% CI [−0.05, 0.79]). EMA and smartphone measurements showed a negligible correlation (r = 0.26, *p* = 0.005, 95% CI [0.10, 0.56]). In contrast, smartphone and bed sensor data did not exhibit a significant correlation (r = 0.17, *p* = 0.108, 95% CI [−0.04, 0.47]), suggesting greater variability in smartphone-based sleep estimates.

**Figure 7.**
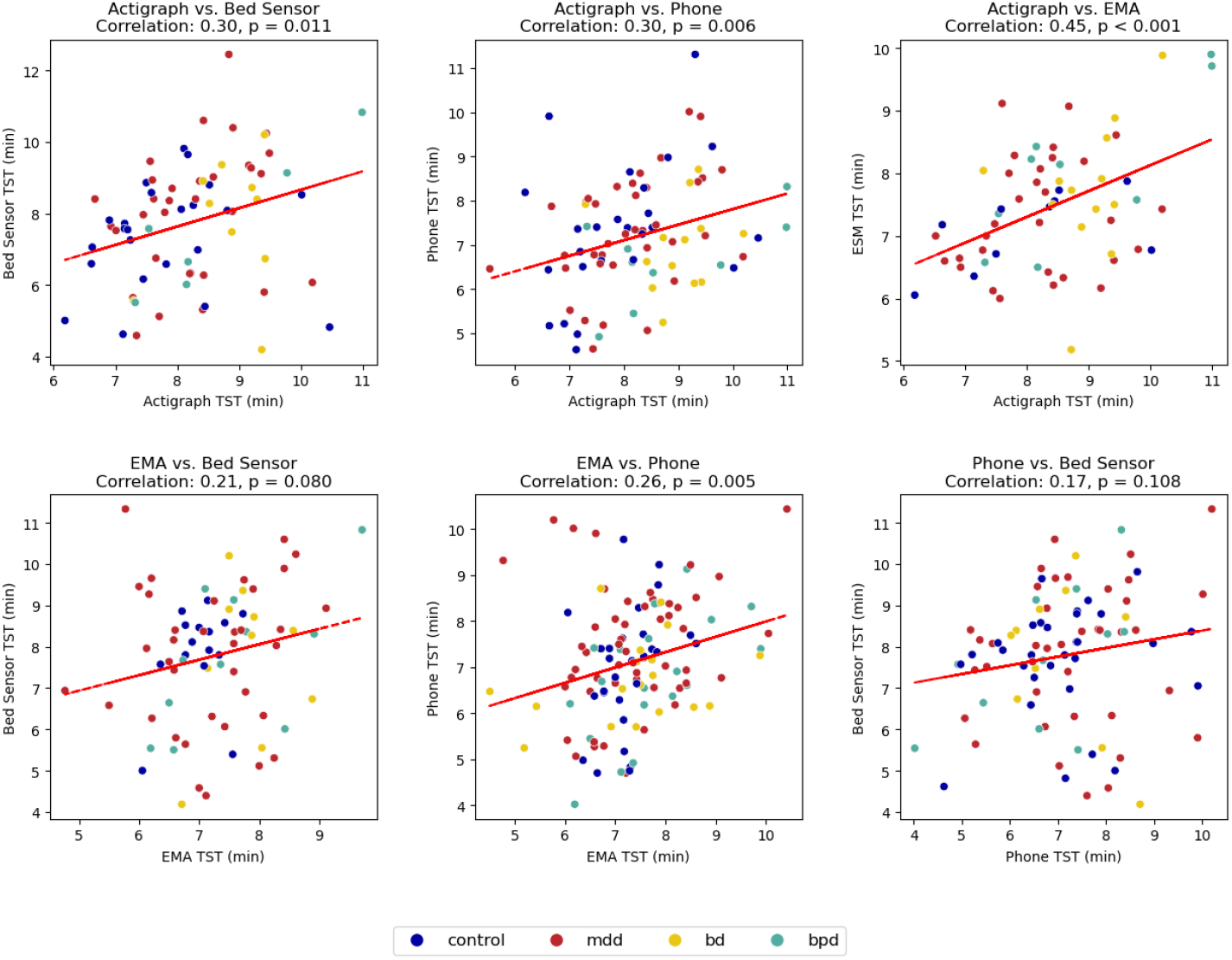
Scatter plots comparing TST across different modalities. All comparisons show weak positive correlations, with EMA vs. bed sensor and smartphone vs. bed sensor not exhibiting statistically significant correlations.

Mean biases in onset time are not statistically significant, whereas offset biases are. TST differences are mostly significant, except for actigraphy vs. bed sensor, ESM vs. bed sensor, and phone vs. bed sensor comparisons.

### 3.3 Factors associated with the alignments of sleep measurements

Factors associated with the alignment of TST, sleep offset, and sleep onset across different modalities are summarized in Table 4, Table 6, and Table 5. Older age was associated with higher alignment in TST between actigraphy and the bed sensor (*β* = *−*0.02, CI [*−*0.04, 0.00], *p* = 0.048), as well as higher alignment in sleep offset between the smartphone and the bed sensor (*β* = *−*0.03, CI [*−*0.06, 0.00], *p* = 0.032). Males exhibited higher alignment compared to females in TST between smartphone and bed sensor measurements (*β* = *−*1.03, CI [*−*1.74, *−*0.33], *p* = 0.004). Longer daylight duration consistently predicted higher alignment in sleep offset for both actigraphy-bed sensor (*β* = *−*0.11, CI [*−*0.22, 0.00], *p* = 0.045) and smartphone-bed sensor comparisons (*β* = −0.11, CI [−0.20, −0.01], *p* = 0.025), highlighting the impact of weather in the alignment between measurement modalities. Patients diagnosed with BD (*β* = 0.89, CI [0.11, 1.68], *p* = 0.026) and BPD (*β* = 0.83, CI [0.01, 1.64], *p* = 0.047) demonstrated a lower alignment in TST between actigraphy and bed sensor compared to controls. No significant predictors of sleep onset alignment were identified.

**Table 4:** TST alignment.

| Total sleep time alignment |  |  |  |  |  |  |  |  |  |
| --- | --- | --- | --- | --- | --- | --- | --- | --- | --- |
|  | Actigraph - Bed |  |  | Phone - Bed |  |  | Actigraph - Phone |  |  |
| *Predictors* | *Estimates* | *CI* | *p* | *Estimates* | *CI* | *p* | *Estimates* | *CI* | *p* |
| age | 0.00 | -0.03 – 0.03 | 0.965 | -0.02 | -0.05 – 0.01 | 0.114 | -0.02 | -0.04 – -0.00 | **0.048** |
| gender | -0.61 | -1.51 – 0.29 | 0.181 | -1.03 | -1.74 – -0.33 | **0.004** | 0.52 | -0.05 – 1.09 | 0.076 |
| MEQ | -0.04 | -0.12 – 0.03 | 0.228 | -0.04 | -0.11 – 0.02 | 0.187 | -0.04 | -0.08 – 0.01 | 0.099 |
| day length (minutes) | -0.09 | -0.19 – 0.02 | 0.103 | -0.04 | -0.12 – 0.05 | 0.366 | -0.07 | -0.14 – -0.01 | **0.026** |
| Group: BD | -0.86 | -2.27 – 0.54 | 0.229 | -0.50 | -1.57 – 0.58 | 0.365 | 0.89 | 0.11 – 1.68 | **0.026** |
| Group: BPD | 0.00 | -1.34 – 1.35 | 0.995 | -0.03 | -1.15 – 1.08 | 0.953 | 0.83 | 0.01 – 1.64 | **0.047** |
| Group: MDD | -0.08 | -1.21 – 1.06 | 0.896 | -0.60 | -1.52 – 0.32 | 0.199 | -0.16 | -0.77 – 0.46 | 0.622 |
| **Random Effects** |  |  |  |  |  |  |  |  |  |
| $\sigma^2$ | 2.47 | | | 2.84 | | | 2.86 | | |
| $\tau^2_0$ | 1.27 user | | | 0.82 user | | | 0.74 user | | |
| ICC | 0.34 |  |  | 0.22 |  |  | 0.21 |  |  |
| N | 49 user |  |  | 52 user |  |  | 77 user |  |  |
| Observations | 891 |  |  | 778 |  |  | 1506 |  |  |
| Marginal R2 / Conditional R2 | 0.045 / 0.369 |  |  | 0.073 / 0.281 |  |  | 0.111 / 0.294 |  |  |

Older age and longer day length were linked to higher TST alignment between actigraphy and phone. Males showed higher alignment than females in TST between phone and bed sensors. Patients with BD and BPD had lower TST alignment between actigraphy and phone compared to healthy controls.

**Table 5:**
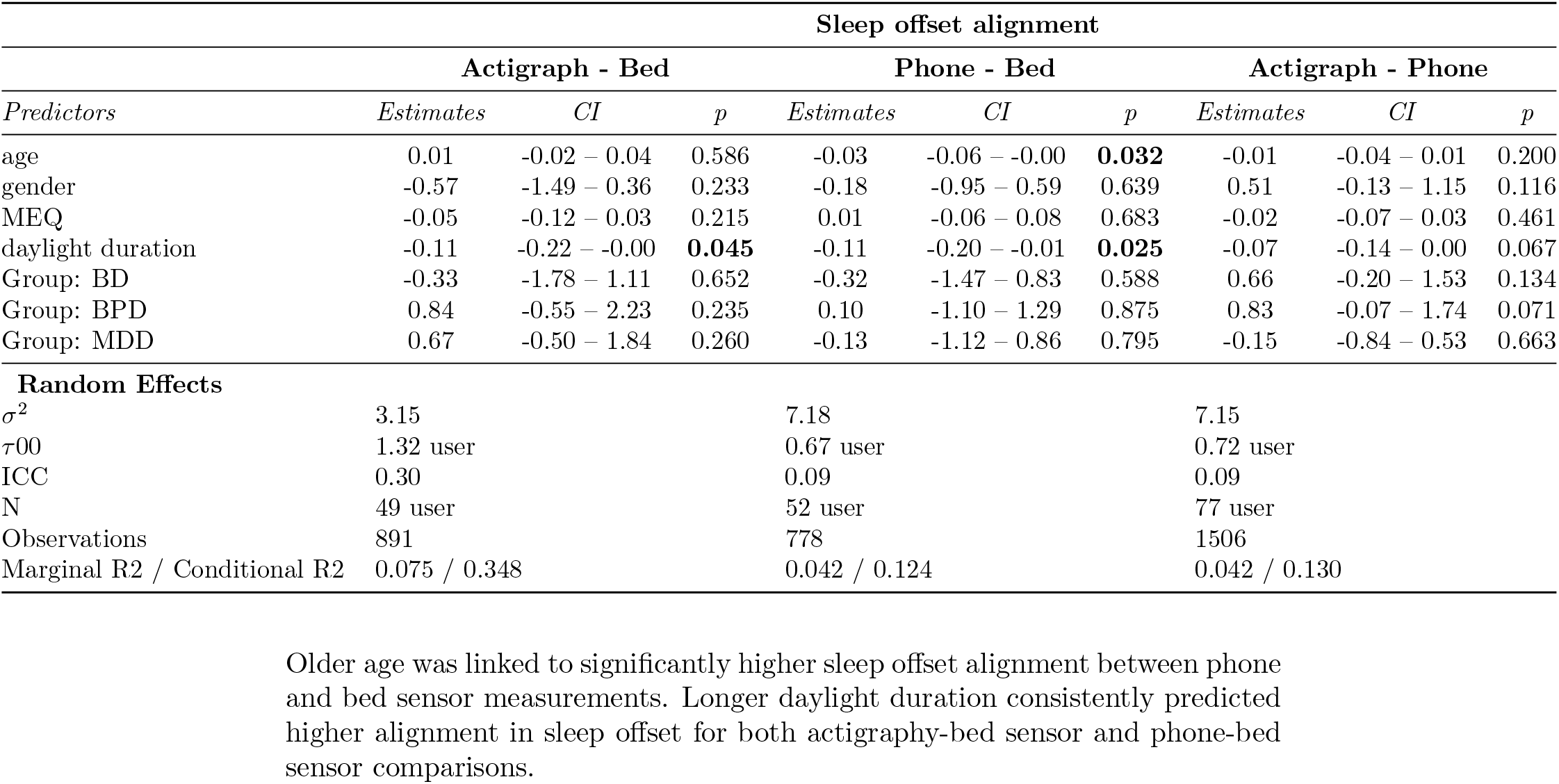
Sleep offset alignment.

**Table 6:**
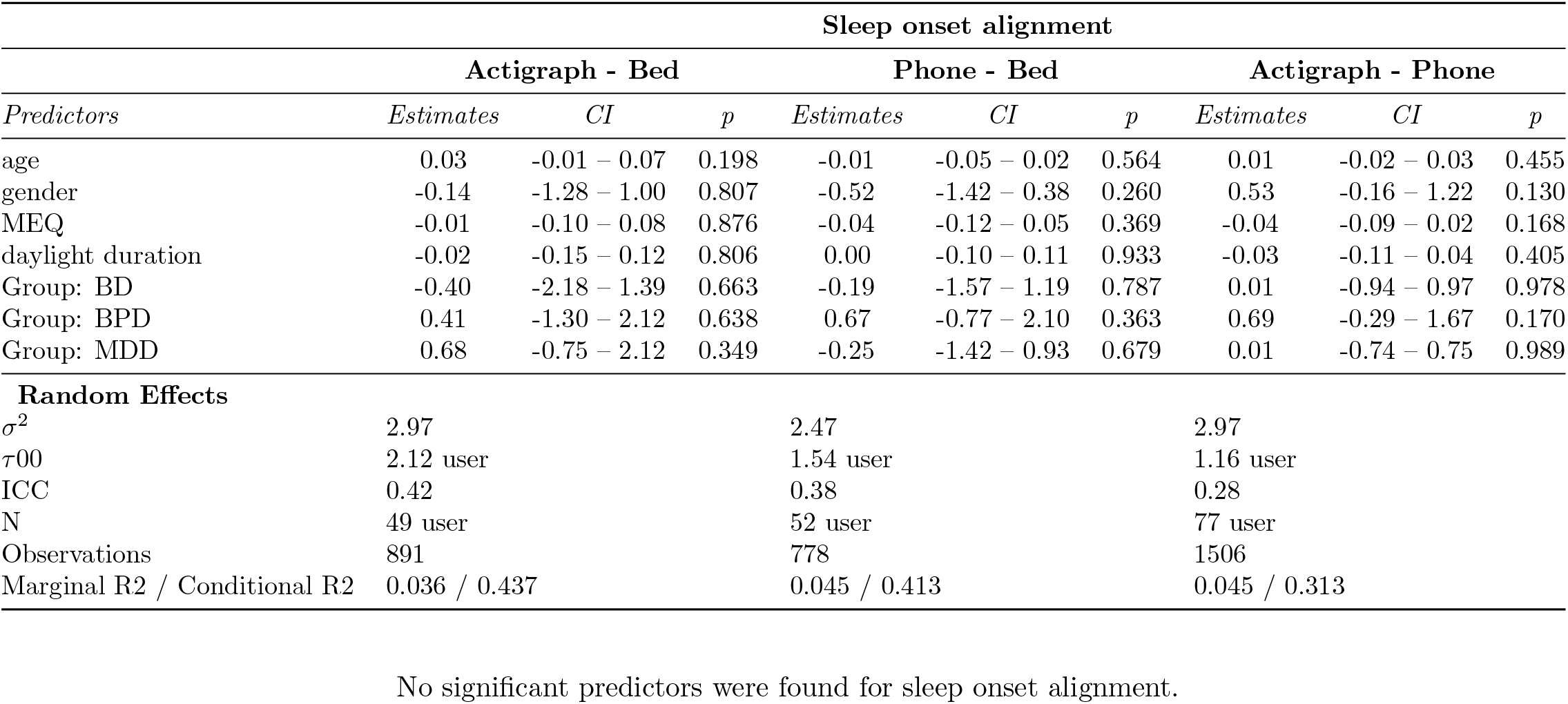
Sleep onset alignment.

## 4 Discussion

Our results showed that patients exhibited greater variability in sleep onset, offset, and TST compared to healthy controls. While actigraphy yielded higher TST, smartphones typically reported lower values. Sleep onset and offset showed stronger correlations across modalities, whereas TST correlations were weaker. Further analysis with linear mixed models revealed that older individuals had better alignment between modalities, while females and patients with BD or BPD showed poorer alignment. Additionally, longer daylight duration was linked to improved alignment in both sleep offset and TST.

Accurate sleep monitoring outside of laboratory settings is increasingly relevant for mental health research, particularly in the absence of PSG as ground truth.^3^ As evidenced in this study, sleep measurements among individuals with depressive disorders exhibited greater between-individual variability compared to the control group, as reflected by the wider confidence intervals. Patients with depression may experience either insomnia, such as trouble falling asleep, frequent awakenings, or early morning awakenings, or hypersomnia. Hypersomnia is often linked to atypical depression, a subtype of major depressive episodes identified in the DSM-5,^51^ and contributes significantly to the variation in sleep patterns among individuals with depression. These findings align with prior research, including Ho et al.,^31^ which reported notable disturbances in both day and night activities among individuals experiencing MDD or symptoms of depression. Their meta-analysis review highlighted disruptions in sleep onset and sleep continuity, poorer sleep efficiency, and greater variability in circadian sleep-wake patterns in relation to healthy controls.

Our results reveal systematic biases in sleep estimation in all measurement modalities. The higher values of TST as measured by actigraphy may be attributed to its limited ability to detect wakefulness during sleep, leading to an overestimation of TST compared to other modalities, a finding consistent with previous studies that have shown similar patterns of overestimation by actigraphy.^13, 52^ On the other hand, smartphone data may have limitations in accurately capturing sleep onset and offset times. The Bland-Altman plots showed that the differences in sleep onset between the smartphone and other modalities were significantly larger, which could contribute to the smartphone’s underestimation of TST. This aligns with findings from Ciman and Wac,^53^ who noted that smartphone algorithms, while more accurate in estimating TST, struggle to detect precise sleep onset and offset. They noted that individuals often stay in bed after waking up before using their smartphones, making it difficult for the smartphone to detect the precise onset time. Similarly, smartphone use before sleep may delay the detected sleep onset. Moreover, the smartphone’s underestimation of TST in our study is consistent with previous research, such as Natale et al.,^54^ which found that smartphone accelerometers generally underestimated TST in healthy individuals, particularly during longer TSTs.

Our findings indicate that smartphone-based measurements exhibited the lowest mean bias and a positive correlation of TST with EMA data. These results partly align with those of Maynard et al.,^55^ who found that a smartphone application strongly correlated with self-reported sleep diaries. However, their study reported that the smartphone overestimated TST compared to actigraphy in healthy adults, whereas our results showed that actigraphy consistently reported higher TST. This discrepancy may stem from methodological differences, as Maynard’s study used a proprietary algorithm to convert accelerometer data into sleep metrics, while our study relied on a simpler approach based on smartphone screen activity events tracking. Additionally, Maynard’s study focused on healthy adults, whereas our study involved psychiatric patients, which may account for the different findings. Given the limited number of studies comparing smartphone-based sleep measurements, Maynard’s study represents the closest methodological comparison. This further highlights the novelty of our study and enhances the validity of our results by showing that smartphone-based sleep measurements can provide meaningful insights even in clinical populations.

Regarding psychiatric populations, Staples et al.^56^ conducted one of the few studies to compare smartphone-based sleep monitoring in individuals with schizophrenia. They found that smartphone accelerometer estimates of TST were moderately correlated with self-reported TST (r = 0.69). However, their study did not compare smartphone estimates directly with actigraphy or other modalities, making it difficult to fully interpret their findings. Our study, which directly compared smartphone-derived TST estimates with those from other objective modalities in psychiatric populations, contributes to the sparse literature on this topic.^20^ We emphasize the necessity for additional research to gain a deeper understanding of the accuracy of smartphone sleep monitoring in clinical populations, particularly when compared to other objective modalities.

Furthermore, the weak correlation in TST across different methods suggests that each device captures distinct aspects of sleep, leading to measurement inconsistencies. Such direct comparisons across multiple modalities within patient groups, without using PSG as a ground truth, are rarely conducted, making this study a unique contribution. One of the few studies in this area is Piantino et al.,^57^ which identified a strong relationship between a bed sensor and an actigraphy device for metrics such as bed entry time, sleep onset, sleep offset, bed exit time, and TST in healthy individuals. However, Piantino et al. also acknowledged that the consistency of the two modalities could be reduced in specific groups, such as individuals with depression or sleep disorders, who may be more likely to lie motionless in bed without achieving sleep. Additionally, in our study, some psychiatric patients were recruited from psychiatric units rather than primary care, where individuals often present with more severe symptoms. As a result, they may be more likely to be overmedicated. Sedative medications can also have muscle-relaxing effects, which may contribute to this phenomenon.

Demographic factors such as age and gender were associated with sleep measurement alignment. Older individuals exhibited better alignment between smartphone and actigraphy estimates, possibly due to more stable sleep patterns. Another explanation could be the reduced smartphone use before bedtime by older participants,^58^ which may align with actigraphy’s overestimation tendencies. This also aligns with findings from Jonasdottir et al.,^59^ who analysed a large global dataset of over 11 million nights from wearable devices across 47 countries. Their study confirmed that older adults exhibit shorter TST, earlier sleep timing, and lower intra-individual variability compared to younger adults, which could contribute to more consistent sleep measurement alignment. Gender differences also played a role, as women showed more discrepancies between smartphone and bed sensor measurements. Jonasdottir et al.^59^ also found that women experience more frequent nighttime awakenings than men, particularly in early to middle adulthood. This suggests that increased sleep fragmentation in women may lead to greater inconsistencies between sensor-based measurements.

Additionally, day length appeared to correlate with better alignment, likely because extended daylight promotes more consistent sleep-wake schedules. These findings align with prior research suggesting that external cues, such as light exposure, play a significant role in regulating sleep timing.^29–31^ This is particularly important given that the study was conducted in Finland, a country prone to extreme fluctuations in daylight. Such fluctuations may have a substantial impact on sleep patterns, making it critical to study and track these changes closely to better understand their effects on sleep and circadian rhythms.

Lastly, the greater alignment observed in BD and BPD patients reinforces the established link between psychiatric disorders and sleep disturbances. Our findings also emphasize the comorbidity between these conditions, consistent with previous literature.^60, 61^ This is also supported by research linking these conditions to irregular sleep patterns. For instance, a meta-analysis found significant variations between BPD patients and healthy participants in sleep stability and architecture indicators, including sleep efficiency, sleep onset delay, TST, and rapid eye movement (REM) sleep parameters.^12^ Similarly, studies examining BD have highlighted disruptions in both quantity and quality, which are often influenced by the manic and depressive episodes characteristic of the disorder.^62^

When using sleep monitoring to assess clinical status or detect early changes, it is important to recognize that there is no definitive standard for directly comparing different modalities. However, we can compare TST with subjective reports from EMA. Although no single device can be definitively identified as the best, we can identify which devices align closely with subjective reports. Our findings showed that, in terms of bias, smartphone had the lowest bias compared to EMA, although the difference was not statistically significant and may be due to random measurement error. Following this, the bed sensor exhibited the next lowest bias, while the actigraph had the highest bias. In contrast, when considering correlation, the actigraph and EMA showed the strongest alignment, suggesting that actigraphy may show a more consistent correlation with subjective sleep reports. However, because actigraphy has been shown to overestimate TST, it is possible that individuals also tend to overestimate their own TST. Clinicians should take these nuances into account, along with the various factors that influence agreement between devices, when choosing a method for sleep monitoring.

A key strength of this study is its naturalistic approach, allowing for real-world sleep monitoring without interfering with participants’ routines. This enhances ecological validity and provides insights into sleep measurement in everyday settings, particularly for individuals with depressive disorders. Additionally, using multiple modalities allows for a comprehensive comparison of different methodologies, highlighting their relative strengths and weaknesses. However, there are important limitations to consider. Our study did not include PSG or sleep logs, as these could have disrupted participants’ natural sleep patterns. Additionally, incorporating PSG would have posed significant challenges, particularly due to the need for frequent lab visits, which could have increased participant burden and potentially led to higher dropout rates, especially among patients. While this approach improves feasibility, it also means that we lack a clinical gold standard for validation. Without PSG data, determining the absolute accuracy of each device remains challenging. Additionally, although EMA provides subjective estimates of TST that align with some objective modalities, it remains susceptible to sleep misperception, where individuals may misjudge their sleep quality or duration.^22^ Other limitations of our study include a relatively small sample size and short follow-up period, which may impact the generalizability of the results. Additionally, the sleep/wake labels used in the study were generated by proprietary algorithms provided by the manufacturers, which may limit the accuracy and comparability of the sleep data. Moreover, differences in sleep patterns may be partly attributable to the use of sleep medication, which is common among psychiatric patients but not typically used by healthy participants. TST outside the 3–13 hour range were excluded based on standard guidelines, which are suitable for healthy individuals but may overlook cases of hypersomnia in patients with depression who may sleep more than 13 hours. Finally, the study was conducted in a single location, resulting in a lack of population diversity. These limitations should be considered when interpreting the results, particularly in clinical contexts.

This study highlights the feasibility of using actigraphy, smartphone data, and bed sensors for sleep tracking in naturalistic settings for patients with depressive disorders. However, it also reveals systematic biases in sleep offset across these modalities, which contribute to discrepancies in TST. Furthermore, our findings highlight that factors such as age, sex, clinical diagnosis, and daylight duration can influence the alignment of sleep measurements across different modalities. This is particularly important in psychiatric populations, where accurate and reliable sleep tracking is essential for research and clinical care.

## Data Availability

Due to the sensitive nature of the data, it is not publicly accessible

## Notes

### Competing Interest Statement

The authors have declared no competing interest.

### Funding Statement

This study did not receive any funding

### Author Declarations

The study was approved by the ethics committee of the Helsinki and Uusimaa Hospital District (HUS), and was granted research permit by HUS Psychiatry.

